# COVID-19 mortality risk correlates inversely with vitamin D3 status, and a mortality rate close to zero could theoretically be achieved at 50 ng/ml 25(OH)D3: Results of a systematic review and meta-analysis

**DOI:** 10.1101/2021.09.22.21263977

**Authors:** Lorenz Borsche, Bernd Glauner, Julian von Mendel

**Author notes:** **Corresponding author:** L.B.

## Abstract

**Background:** Much research shows that blood calcidiol (25(OH)D3) levels correlate strongly with SARS-CoV-2 infection severity. There is open discussion regarding whether low D3 is caused by the infection or if deficiency negatively affects immune defense. The aim of this study was to collect further evidence on this topic.

**Methods:** Systematic literature search was performed to identify retrospective cohort as well as clinical studies on COVID-19 mortality rates versus D3 blood levels. Mortality rates from clinical studies were corrected for age, sex and diabetes. Data were analyzed using correlation and linear regression.

**Results:** One population study and seven clinical studies were identified, which reported D3 blood levels pre-infection or on the day of hospital admission. They independently showed a negative Pearson correlation of D3 levels and mortality risk (r(17)=-.4154, p=.0770/r(13)=-.4886, p=.0646). For the combined data, median (IQR) D3 levels were 23.2 ng/ml (17.4 – 26.8), and a significant Pearson correlation was observed (r(32)=-.3989, p=.0194). Regression suggested a theoretical point of zero mortality at approximately 50 ng/ml D3.

**Conclusions:** The two datasets provide strong evidence that low D3 is a predictor rather than a side effect of the infection. Despite ongoing vaccinations, we recommend raising serum 25(OH)D levels to above 50 ng/ml to prevent or mitigate new outbreaks due to escape mutations or decreasing antibody activity.

**Trial registration:** Not applicable.

## Background

The SARS-CoV-2 pandemic causing acute respiratory distress syndrome (ARDS) has lasted for more than 18 months. It has created a major global health crisis due to the high number of patients requiring intensive care, and the high death rate has substantially affected everyday life through contact restrictions and lockdowns. According to many scientists and medical professionals, we are far from the end of this disaster and hence must learn to coexist with the virus for several more years, perhaps decades [1,2].

It is realistic to assume that there will be new mutations, which are possibly more infectious or more deadly. In the known history of virus infections, we have never faced a similar global spread. Due to the great number of viral genome replications that occur in infected individuals and the error-prone nature of RNA-dependent RNA polymerase, progressive accrual mutations do and will continue to occur [3–5]. Thus, similar to other virus infections such as influenza, we have to expect that the effectiveness of vaccination is limited in time, especially with the current vaccines designed to trigger an immunological response against a single viral protein [6–8].

We have already learned that even fully vaccinated people can be infected [9]. Currently, most of these infections do not result in hospitalization, especially for young individuals without comorbidities. However, these infections are the basis for the ongoing dissemination of the virus in a situation where worldwide herd immunity against SARS-CoV-2 is rather unlikely. Instead, humanity could be trapped in an insuperable race between new mutations and new vaccines, with an increasing risk of newly arising mutations becoming resistant to the current vaccines [3,10,11]. Thus, a return to normal life in the near future seems unlikely. Mask requirements as well as limitations of public life will likely accompany us for a long time if we are not able to establish additional methods that reduce virus dissemination.

Vaccination is an important part in the fight against SARS-CoV-2 but, with respect to the situation described above, should not be the only focus. One strong pillar in the protection against any type of virus infection is the strength of our immune system [12]. Unfortunately, thus far, this unquestioned basic principle of nature has been more or less neglected by the responsible authorities. It is well known that our modern lifestyle is far from optimal with respect to nutrition, physical fitness and recreation. In particular, many people are not spending enough time outside in the sun, even in summer. The consequence is widespread vitamin D deficiency, which limits the performance of their immune systems, resulting in the increased spread of some preventable diseases of civilization, reduced protection against infections and reduced effectiveness of vaccination [13].

In this publication, we will demonstrate that vitamin D3 deficiency, which is a well-documented worldwide problem [13–19,179], is one of the main reasons for severe courses of SARS-CoV-2 infections. The fatality rates correlate well with the findings that elderly people, black people and people with comorbidities show very low vitamin D3 levels [16,20–22]. Additionally, with only a few exceptions, we are facing the highest infection rates in the winter months and in northern countries, which are known to suffer from low vitamin D3 levels due to low endogenous sun-triggered vitamin D3 synthesis [23–26].

Vitamin D3 was first discovered at the beginning of the 19^th^ century as an essential factor needed to guarantee skeletal health. This discovery came after a long period of dealing with the dire consequences of rickets, which causes osteomalacia (softening of bones). This disease especially affected children in northern countries, who were deprived of sunlight and often worked in dark production halls during the industrial revolution [27]. At the beginning of the 20^th^ century, it became clear that sunlight can cure rickets by triggering vitamin D3 synthesis in the skin. Cod liver oil is recognized as a natural source of vitamin D3 [28]. At the time, a blood level of 20 ng/ml was sufficient to stop osteomalacia. This target is still the recommended blood level today, as stated in many official documents [29]. In accordance with many other publications, we will show that this level is considerably too low to guarantee optimal functioning of the human body.

In the late 1920s, Adolf Windaus elucidated the structure of vitamin D3. The metabolic pathway of vitamin D3 (biochemical name cholecalciferol) is shown in Figure 1 [30]. The precursor, 7-dehydrocholesterol, is transformed into cholecalciferol in our skin by photoisomerization caused by UV-B exposure (wavelength 280–315 nm). After transportation to the liver, cholecalciferol is hydroxylated, resulting in 25-hydroxycholecalciferol (25(OH)D3, also called calcidiol), which can be stored in fat tissue for several months and is released back into blood circulation when needed. The biologically active form is generated by a further hydroxylation step, resulting in 1,25-dihydroxycholecalciferol (1,25(OH)_2_D3, also called calcitriol). Early investigations assumed that this transformation takes place mainly in the kidney.

**Fig. 1.**
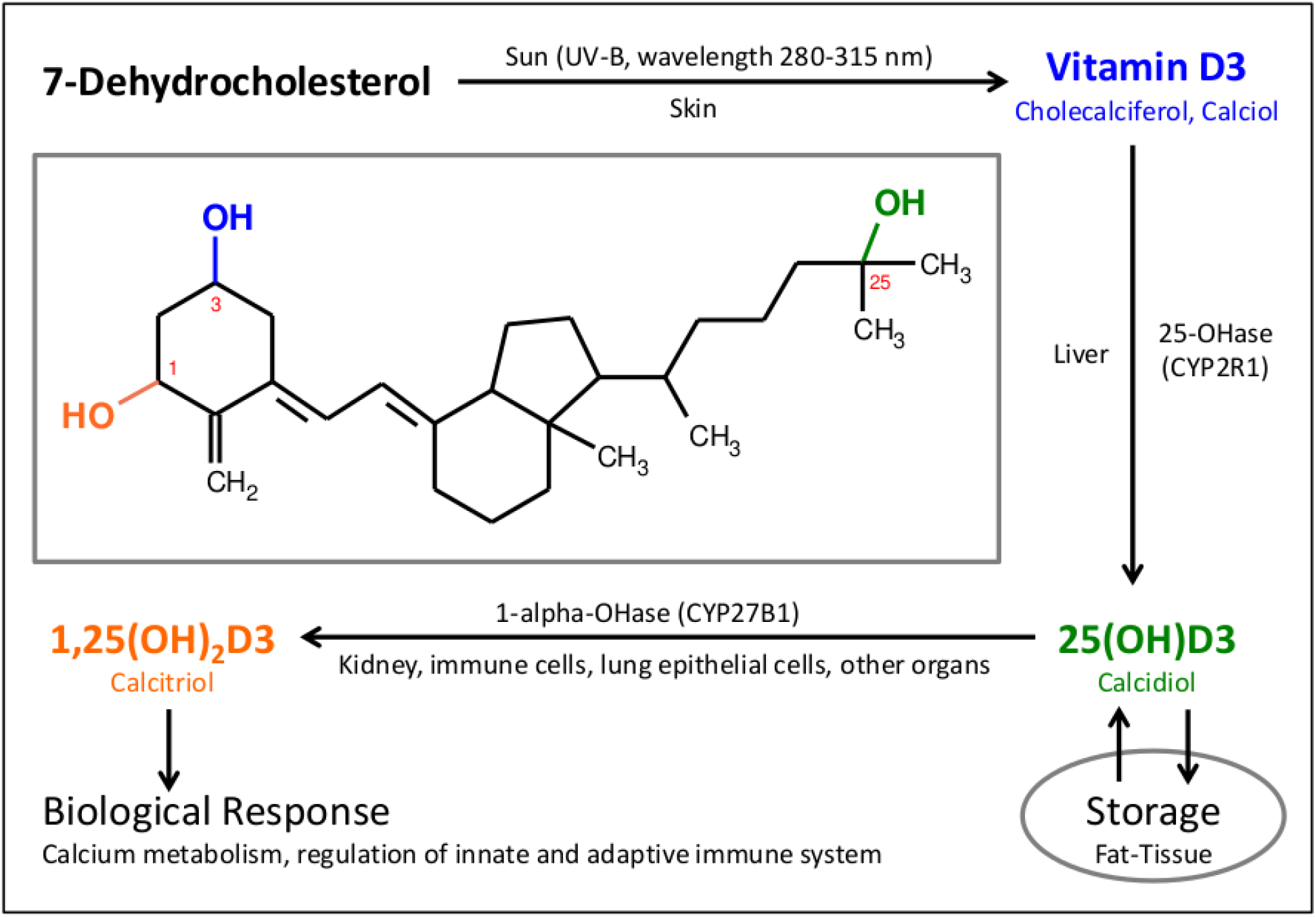
Metabolic Pathway of Vitamin D3. The vitamin D pathway is characterized by two subsequent hydroxylation steps. In the liver, 25-Hydroxylase produces 25(OH)D3 (calcidiol), which can be stored in fat tissue. 1-Alpha-hydroxylase generates the active steroid hormone 1,25(OH)2D3 (calcitriol), which regulates calcium metabolism as well as the innate and adaptive immune system.

Over the last decades, knowledge regarding the mechanisms through which vitamin D3 affects human health has improved dramatically. It was discovered that the vitamin D3 receptor (VDR) and the vitamin D3 activating enzyme 1-α-hydroxylase (CYP27B1) are expressed in many cell types that are not involved in bone and mineral metabolism, such as the intestine, pancreas, and prostate as well as cells of the immune system [31–35]. This finding demonstrates the important, much wider impact of vitamin D3 on human health than previously understood [36,37]. Vitamin D turned out to be a powerful epigenetic regulator, influencing more than 2500 genes [38] and impacting dozens of our most serious health challenges [39], including cancer [40,41], diabetes mellitus [42], acute respiratory tract infections [43], chronic inflammatory diseases [44] and autoimmune diseases such as multiple sclerosis [45].

In the field of human immunology, the extrarenal synthesis of the active metabolite calcitriol-1,25(OH)_2_D3-by immune cells and lung epithelial cells has been shown to have immunomodulatory properties [46–51]. Today, a compelling body of experimental evidence indicates that activated vitamin D3 plays a fundamental role in regulating both innate and adaptive immune systems [52–55]. Intracellular vitamin D3 receptors (VDRs) are present in nearly all cell types involved in the human immune response, such as monocytes/macrophages, T cells, B cells, natural killer (NK) cells, and dendritic cells (DCs). Receptor binding engages the formation of the “vitamin D3 response element” (VDRE), regulating a large number of target genes involved in the immune response [56]. As a consequence of this knowledge, the scientific community now agrees that calcitriol is much more than a vitamin but rather a highly effective hormone with the same level of importance to human metabolism as other steroid hormones.

The blood level ensuring the reliable effectiveness of vitamin D3 with respect to all its important functions came under discussion again, and it turned out that 40–60 ng/ml is preferable [37], which is considerably above the level required to prevent rickets.

Long before the SARS-CoV-2 pandemic, an increasing number of scientific publications showed the effectiveness of a sufficient vitamin D3 blood level in curing many of the human diseases caused by a weak or unregulated immune system [37,57–59]. This includes all types of virus infections [43,60–68,180], with a main emphasis on lung infections that cause ARDS [69–71], as well as autoimmune diseases [45,62,72,73]. However, routine vitamin D3 testing and supplementation are still not established today. Unfortunately, it seems that the new findings about vitamin D3 have not been well accepted in the medical community. Many official recommendations to define vitamin D3 deficiency still stick to the 20 ng/ml established 100 years ago to cure rickets [74].

Additionally, many recommendations for vitamin D3 supplementation are in the range of 5 to 20 μg per day (200 to 800 international units), which is much too low to guarantee the optimal blood level of 40–60 ng/ml [37,75]. One reason for these incorrect recommendations turned out to be calculation error [76,77]. Another reason for the error is because vitamin D3 treatment to cure osteomalacia was commonly combined with high doses of calcium to support bone calcification. When examining for the side effects of overdoses of such combination products, it turned out that there is a high risk of calcium deposits in blood vessels, especially in the kidney. Today, it is clear that such combination preparations are nonsensical because vitamin D3 stimulates calcium uptake in the intestine itself. Without calcium supplementation, even very high vitamin D3 supplementation does not cause vascular calcification, especially if another important finding is included. Even when calcium blood levels are high, the culprit for undesirable vascular calcification is not vitamin D but insufficient blood levels of vitamin K2. Thus, daily vitamin D3 supplementation in the range of 4000 to 10,000 units (100 to 250 μg) needed to generate an optimal vitamin D3 blood level in the range of 40–60 ng/ml has been shown to be completely safe when combined with approximately 200 μg/ml vitamin K2 [78–80]. However, this knowledge is still not widespread in the medical community, and obsolete warnings about the risks of vitamin D3 overdoses unfortunately are still commonly circulating.

Based on these circumstances, the SARS-CoV-2 pandemic is becoming the second breakthrough in the history of vitamin D3 association with disease (after rickets), and we have to ensure that full advantage is being taken of its medical properties to keep people healthy. The most life-threatening events in the course of a SARS-CoV-2 infection are ARDS and cytokine release syndrome (CRS). It is well established that vitamin D3 is able to inhibit the underlying metabolic pathways [81,82] because a very specific interaction exists between the mechanism of SARS-CoV-2 infection and vitamin D3:

Angiotensin-converting enzyme 2 (ACE2), a part of the renin-angiotensin system (RAS), serves as the major entry point for SARS-CoV-2 into cells (Fig. 2). When SARS-CoV-2 is attached to ACE2 its expression is reduced, thus causing lung injury and pneumonia [83,84,175]. Vitamin D3 is a negative RAS modulator by inhibition of renin expression and stimulation of ACE2 expression. It therefore has a protective role against ARDS caused by SARS-CoV-2. Sufficient vitamin D3 levels prevent the development of ARDS by reducing the levels of angiotensin II and increasing the level of angiotensin-(1,7) [18,85,86,172,173,176].

**Fig. 2.**
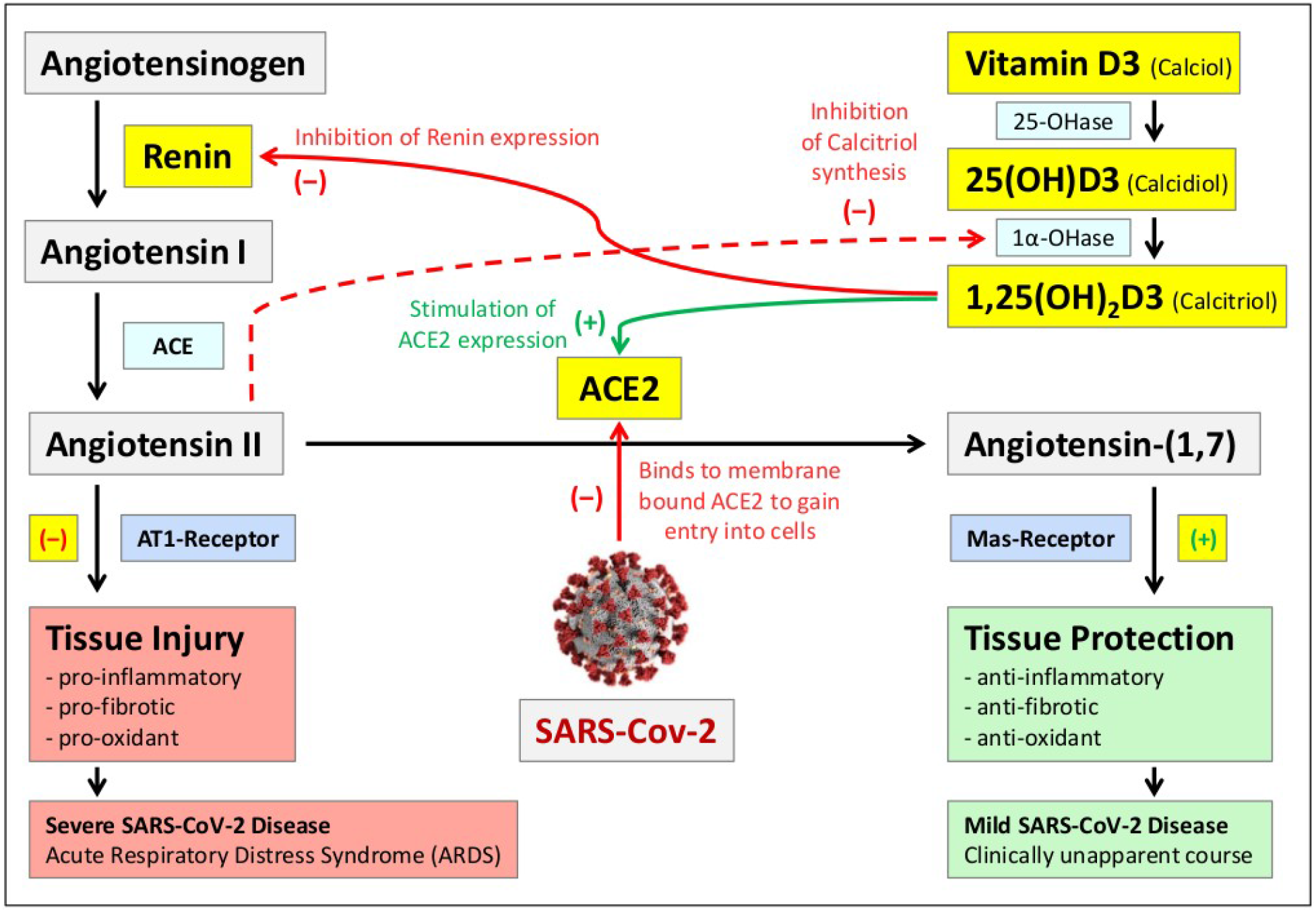
Interaction of Vitamin D3 with the Renin-Angiotensin System (RAS) The renin-angiotensin system (RAS) is an important regulator of blood volume and systemic vascular resistance for the adjustment of blood pressure. The balance between angiotensin II and angiotensin-(1.7) is a critical factor for the proper functioning of the system (175). Angiotensin-converting enzyme 2 (ACE2) is responsible for converting angiotensin II to angiotensin-(1,7). Angiotensin II primarily triggers vasoconstriction but can also cause inflammation, fibrosis and oxidative stress in the absence of its counterpart, angiotensin-(1,7). ACE2 is the primary receptor of SARS-CoV-2, which decreases its activity, leading to an increase in angiotensin II levels and a decrease in angiotensin-(1,7) levels. This effect ultimately triggers SARS-CoV-2-induced “acute respiratory distress syndrome” (ARDS) [83,84]. Calcitriol, the active metabolite of vitamin D3, minimizes this effect by inhibiting renin expression and thus angiotensin II synthesis and by stimulating ACE2 expression [172,173], enhancing the conversion of angiotensin II to angiotensin-(1,7). Thus, insufficient vitamin D blood levels lead to the development of severe courses of SARS-CoV-2 disease. In addition, it has been shown that high angiotensin II levels lead to downregulation of the enzyme 1-alpha-hydroxylase [174], which is required for the formation of calcitriol, thereby exacerbating the negative consequences of vitamin D deficiency.

There are several additional important functions of vitamin D3 supporting immune defense [18,75,87,88]:

- Vitamin D decreases the production of Th1 cells. Thus, it can suppress the progression of inflammation by reducing the generation of inflammatory cytokines [72,89,90].
- Vitamin D3 reduces the severity of cytokine release syndrome (CRS). This “cytokine storm” causes multiple organ damage and is therefore the main cause of death in the late stage of SARS- CoV-2 infection. The systemic inflammatory response due to viral infection is attenuated by promoting the differentiation of regulatory T cells [91–94].
- Vitamin D3 induces the production of the endogenous antimicrobial peptide cathelicidin (LL-37) in macrophages and lung epithelial cells, which acts against invading respiratory viruses by disrupting viral envelopes and altering the viability of host target cells [51,95–100].
- Experimental studies have shown that vitamin D and its metabolites modulate endothelial function and vascular permeability via multiple genomic and extragenomic pathways [101,102].
- Vitamin D reduces coagulation abnormalities in critically ill COVID-19 patients [103–105].

A rapidly increasing number of publications are investigating the vitamin D3 status of SARS-CoV-2 patients and have confirmed both low vitamin D levels in cases of severe courses of infection [106–121] and positive results of vitamin D3 treatments [122–128]. Therefore, many scientists recommend vitamin D3 as an indispensable part of a medical treatment plan to avoid severe courses of SARS-CoV-2 infection [14,18,75,82,129,130], which has additionally resulted in proposals for the consequent supplementation of the whole population [131]. A comprehensive overview and discussion of the current literature is given in a review by Linda Benskin [132]. Unfortunately, all these studies are based on relatively low numbers of patients. Well-accepted, placebo-controlled, double-blinded studies are still missing.

The finding that most SARS-CoV-2 patients admitted to hospitals have vitamin D3 blood levels that are too low is unquestioned even by opponents of vitamin D supplementation. However, there is an ongoing discussion as to whether we are facing a causal relationship or just a decline in the vitamin D levels caused by the infection itself [82,133,134,181].

There are reliable data on the average vitamin D3 levels in the population [15,19,135] in several countries, in parallel to the data about death rates caused by SARS-CoV-2 in these countries [136,137]. Obviously, these vitamin D3 data are not affected by SARS-CoV-2 infections. While meta-studies using such data [25,130,134,138] are already available, our goal was to analyze these data in the same manner as selected clinical data. In this article, we identify a vitamin D threshold that virtually eliminates excess mortality caused by SARS-CoV-2. In contrast to published D3/SARS-CoV-2 correlations [139–141,182-185], our data include studies assessing preinfection vitamin D values as well as studies with vitamin D values measured post-infection latest on the day after hospitalization. Thus, we can expect that the measured vitamin D status is still close to the preinfection level. In contrast to other meta-studies which also included large retrospective cohort studies [184-185], our aim was to perform regressions on the combined data after correcting for patient characteristics.

These results from independent datasets, which include data from before and after the onset of the disease, also further strengthen the assumption of a causal relationship between vitamin D3 blood levels and SARS-CoV-2 death rates. Our results therefore also confirm the importance of establishing vitamin D3 supplementation as a general method to prevent severe courses of SARS-CoV-2 infections.

## Methods

### Search Strategy and Selection Criteria

Initially, a systematic literature review was performed to identify relevant COVID-19 studies. Included studies were observational cohort studies that grouped two or more cohorts by their vitamin D3 values and listed mortality rates for the respective cohorts. PubMed and the https://c19vitamind.com registry were searched according to Table 1. Subsequently, titles and abstracts were screened, and full-text articles were further analyzed for eligibility.

**Table 1.** Search Strategy.

| Source | Search Strategy | Time frame |
| --- | --- | --- |
| PubMed | COVID-19 Search String from [142]<br>AND (“vitamin d” or “d3” or “25(OH)D”<br>or “25-hydroxyvitamin D”) | November 1, 2019 - March<br>27, 2021 |
| https://<br>c19vitamind.com | Restriction to category “Levels” | November 1, 2019 - March<br>27, 2021 |

### Data Analysis

Collected studies were divided into a population study [143] and seven hospital studies. Notably, these data sources are fundamentally different, as one assesses vitamin D values long-term, whereas the other measures vitamin D values postinfection, thereby masking a possible causal relationship between the preinfection vitamin D level and mortality.

Several corrections for the crude mortality rates (CMRs) recorded by Ahmad were attempted to understand the underlying causes within the population study data and the outliers. In the end, none were used in the final data evaluation to avoid the risk of introducing hidden variables that also correlate with D3.

Mortality rates and D3 blood levels from studies on hospitalized COVID-19 patients were assembled in a separate dataset. When no median D3 blood levels were provided for the individual study cohorts, the IQR, mean±SD or estimated values within the grouping criteria were used in that order. Patient characteristics, including age IQR, sex and diabetes status, were used to compute expected mortality rates with a machine learning model [144]. Based on the expected mortality rate, the observed mortality rates were corrected for the specific cohorts.

The two datasets were combined, and the mortality rates of the hospital studies were scaled according to the mortality range of the population studies, resulting in a uniform list of patient cohorts, their vitamin D status and dimensionless mortality coefficients. Linear regressions (OLS) and Pearson and Spearman correlations of vitamin D and the mortality values for the separate and combined datasets were generated with a Python 3.7 kernel using the scipy.stats 1.7.0 and statsmodels 0.12.2 libraries in a https://deepnote.com Jupyter notebook.

## Results

Database and registry searches resulted in 563 and 66 records, respectively. Nonsystematic web searches accounted for 13 studies, from which an additional 31 references were assessed. After removal of 104 duplicates and initial screening, 44 studies remained. Four meta-studies, one comment, one retracted study, one report with unavailable data, one wrong topic report and one Russian language record were excluded. The remaining 35 studies were assessed in full text, 20 of which did not meet the eligibility criteria due to their study design or lack of quantitative mortality data. Four further studies were excluded due to missing data for individual patient cohorts. Finally, three studies were excluded due to skewed or nonrepresentative patient characteristics, as reviewed by LB and JVM [145–147]. Eight eligible studies for quantitative analysis remained, as listed in Table 2. A PRISMA flowchart [148] is presented in Figure 3.

**Table 2.**
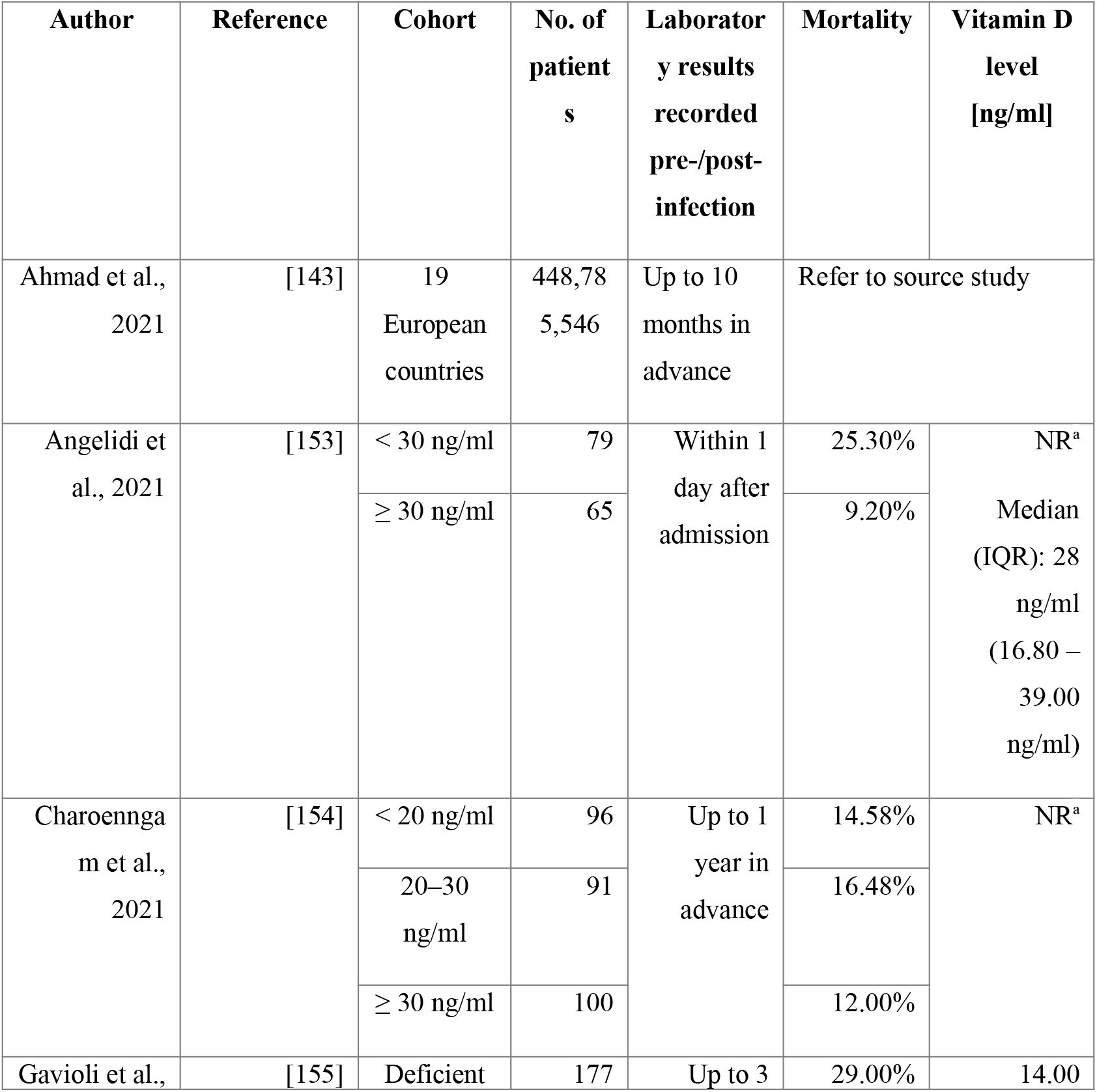
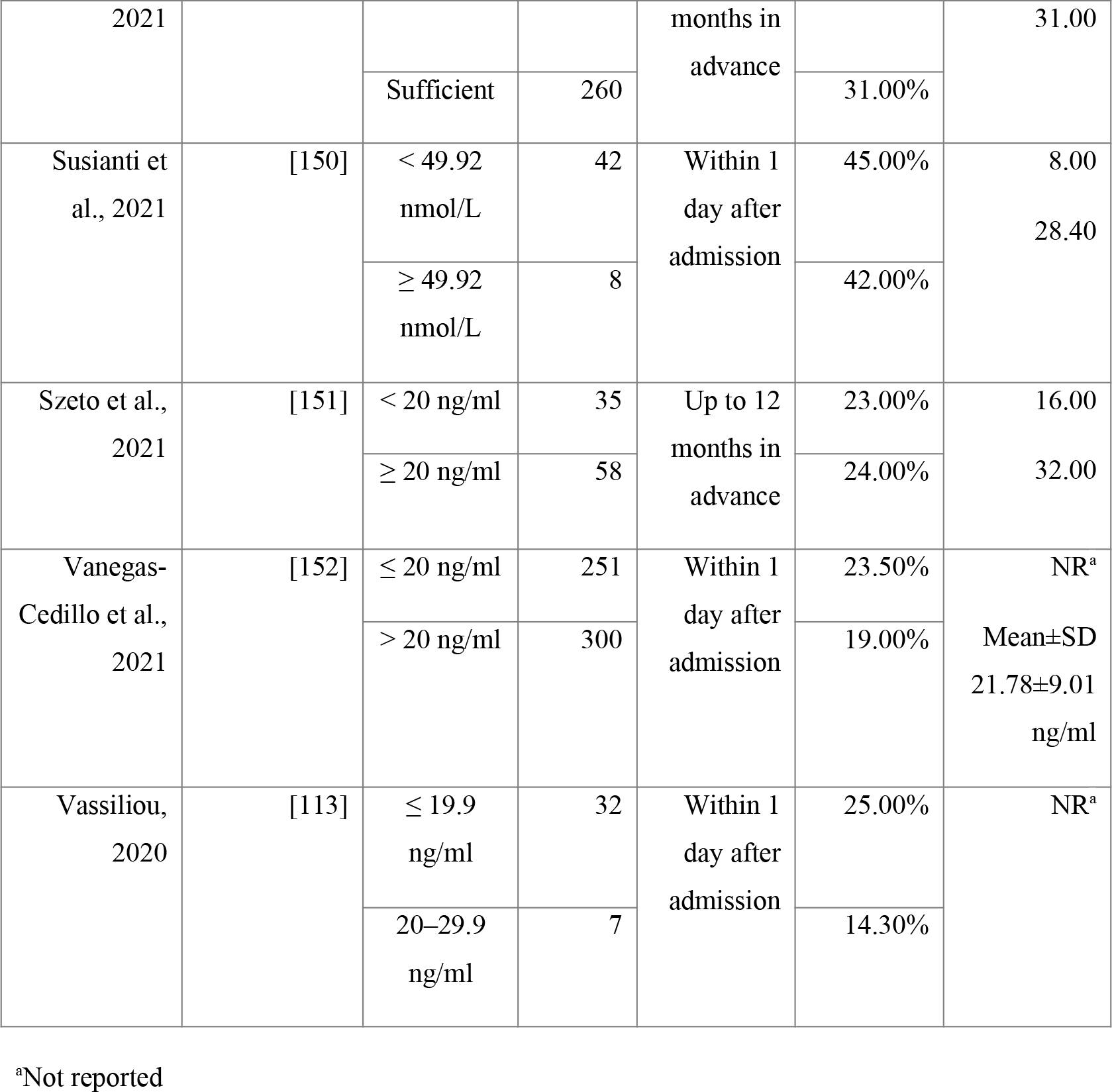
Eligible studies.

**Fig. 3.**
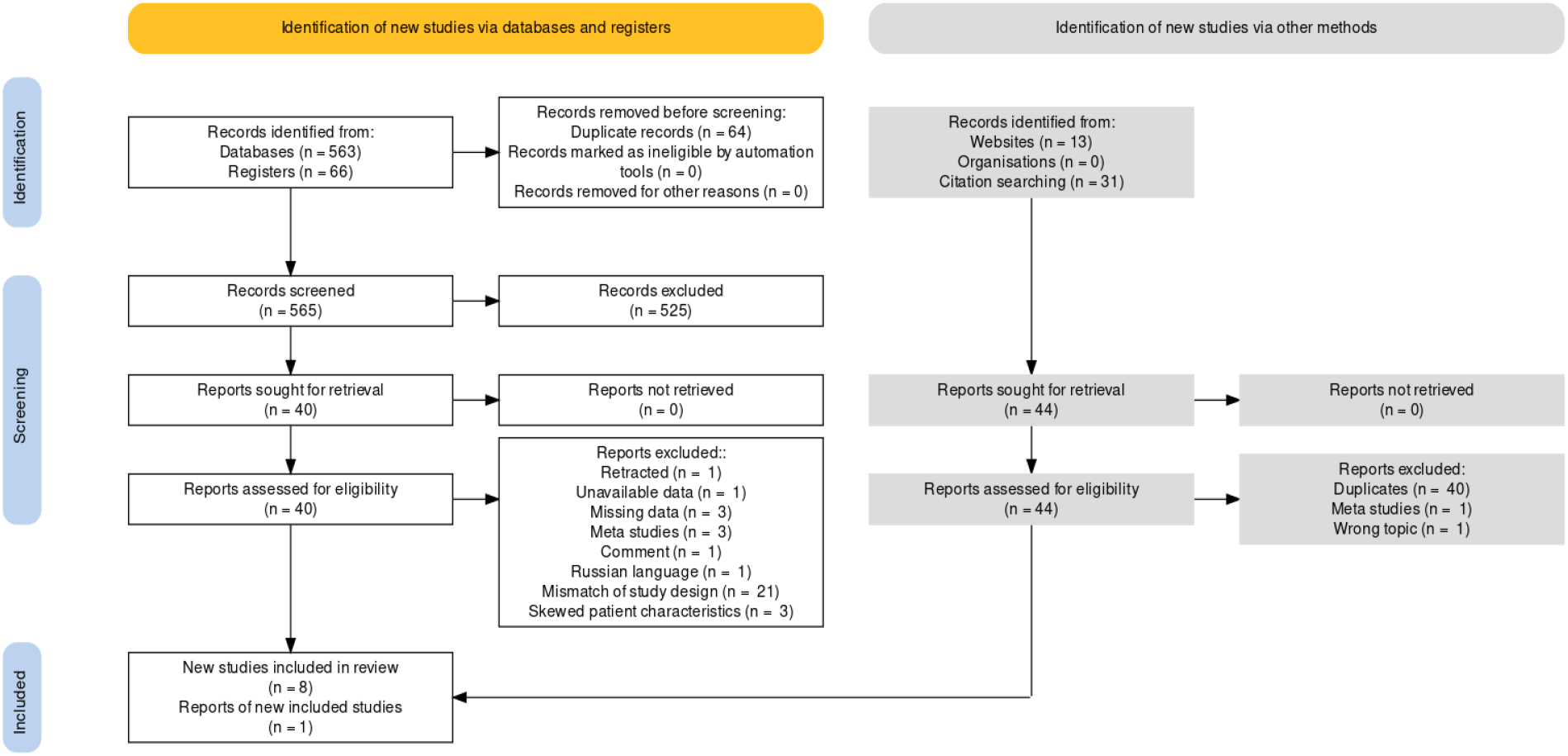
Flowchart of the search strategy and selection process [149].

The observed median (IQR) vitamin D value over all collected study cohorts was 23.2 ng/ml (17.4 – 26.8). A frequency distribution of vitamin D levels is shown in Figure 4.

**Fig. 4.**
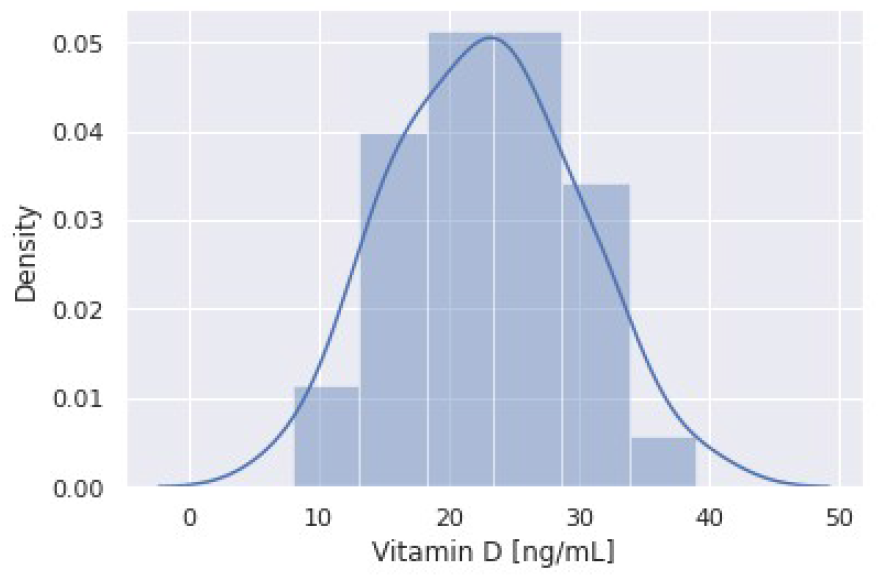
Frequency distribution of vitamin D levels of all evaluated study cohorts.

One population study by Ahmad et al. [143] was identified. Therein, the CMRs are compiled for 19 European countries based on COVID-19 pandemic data from Johns Hopkins University [156] in the time frame from March 21, 2020, to January 22, 2021, as well as D3 blood levels for the respective countries collected by literature review. Furthermore, the proportions of the 70+ age population were collected. The median vitamin D3 level across countries was 23.2 ng/ml (19.9 – 25.5 ng/ml). A moderately negative Spearman’s correlation with the corresponding mean vitamin D3 levels in the respective populations was observed at r_s_=-.430 (95% CI: -.805 – -.081). No further adjustments of these CMR values were performed by Ahmad. The correlations shown in Table 3 suggest the sex/age distribution, diabetes and the rigidity of public health measures as some of the causes for outliers within the Ahmad dataset. However, this has little effect on the further results discussed below.

**Table 3.** Attempted corrections of the CMR values in the population study by Ahmad.

| Method | Reference | Resulting Pearson correlation CMR ~ D3 |
| --- | --- | --- |
| None | – | $r(17) = -.4154$, $p = .0770$ |
| Two most extreme outliers removed | – | $r(15) = -.3471$, $p = .1722$ |
| Rigidity of public health measures | [157] | $r(17) = -.4662$, $p = .0442$ |
| Sex/age distribution, diabetes | [158,159] | $r(17) = -.5113$, $p = .0253$ |
| Expected SARS-COV-2 positive rate for given D3 level | [115] | $r(17) = -.5997$, $p = .0066$ |

The extracted data from seven hospital studies showed a median vitamin D3 level of 23.2 ng/ml (14.5 – 30.9 ng/ml). These data are plotted after correction of patient characteristics and scaling in combination with the data points from Ahmad in Figure 5.

**Fig. 5.**
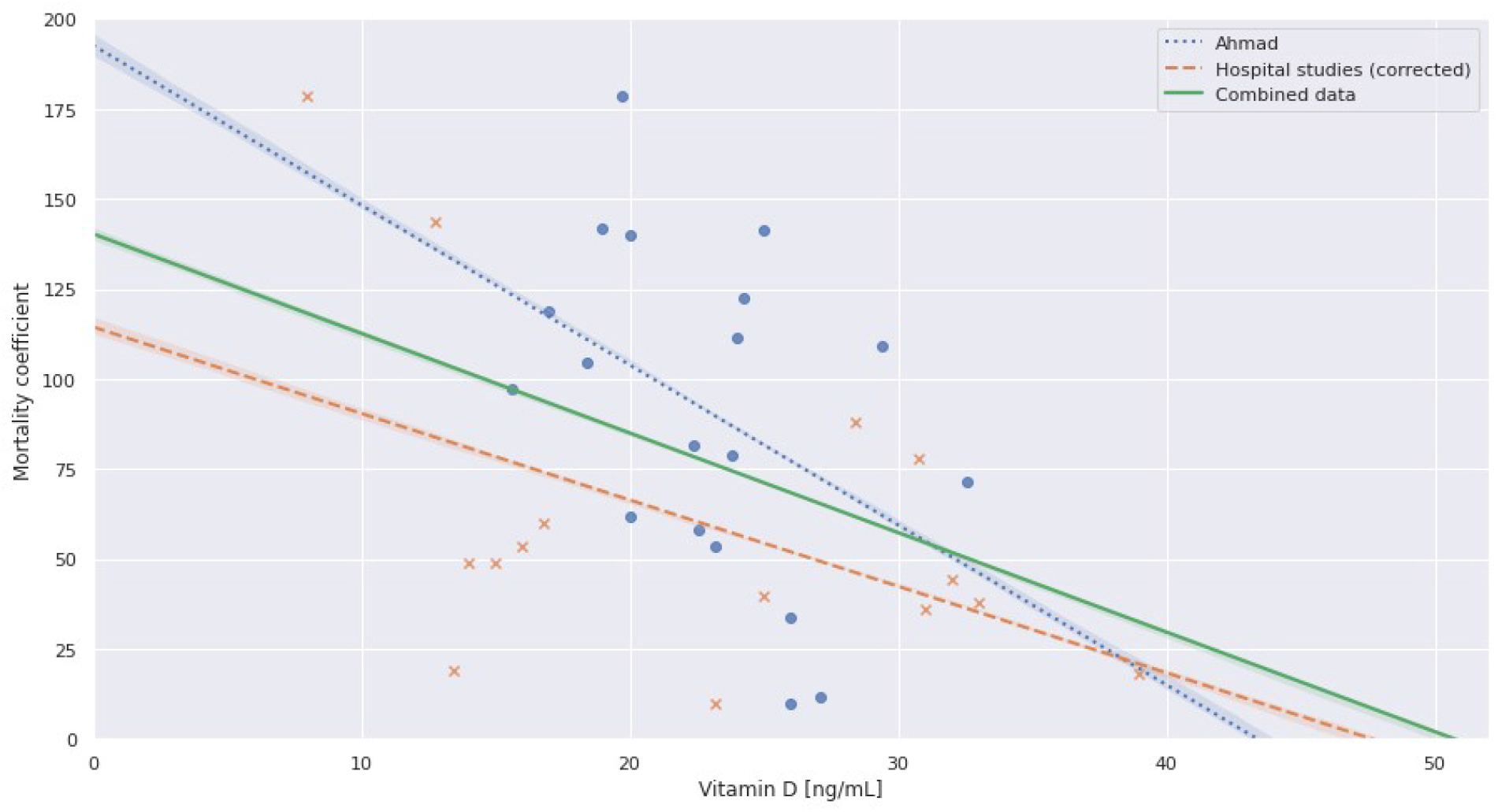
Scatter plot and OLS regressions of the individual and combined datasets.

The correlation results are shown in Table 4 in which the combined data show a significant negative Pearson correlation at r(32)=-.3989, p=.0194. The linear regression results can be found in Table 5. The regression for the combined data intersects the D3 axis at 50.7 ng/ml, suggesting a theoretical point of zero mortality.

**Table 4.** Correlation of mortality and vitamin D blood levels for the respective datasets.

|  | Ahmad | Hospital studies<br>(corrected) | Combined |
| --- | --- | --- | --- |
| **Pearson**<br>**correlation**<br>**(Mortality~Vit D)** | $r(17)=-.4154$,<br>$p=.0770$ | $r(13)=-.4886$, $p=.0646$ | $r(32)=-.3989$,<br>$p=.0194$ |
| **Spearman**<br>**correlation**<br>**(Mortality~Vit D)** | $r_s=-.4300$, $p=.0661$,<br>$N=19$ | $r_s=-.469$, $p=.0786$, $N=15$ | $r_s=-.3698$,<br>$p=.03136$, $N=34$ |

**Table 5.** OLS regressions for the respective datasets.

|  | Ahmad | Hospital studies<br>(corrected) | Combined |
| --- | --- | --- | --- |
| **Intercept** | 192.6788 | 114.4156 | 140.2880 |
| **Coefficient** | -4.4408 | -2.4015 | -2.7654 |
| **R<sup>2</sup>** | .173 | .239 | .159 |
| **Adj. R<sup>2</sup>** | .124 | .180 | .133 |
| **Prob (F-Statistic)** | .0770 | .0646 | .0194 |
| **AIC** | 198.7 | 156.5 | 356.8 |
| **BIC** | 200.6 | 158.0 | 359.8 |
| **Prob (Omnibus)** | .342 | .568 | .436 |
| **Durbin-Watson** | 1.238 | 1.514 | 1.217 |
| **Prob (Jarque-Bera)** | .591 | .662 | .572 |

## Discussion

This study illustrates that, at a time when vaccination was not yet available, patients with sufficiently high D3 serum levels preceding the infection were highly unlikely to suffer a fatal outcome. The partial risk at this D3 level seems to vanish under the normal statistical mortality risk for a given age and in light of given comorbidities. This correlation should have been good news when vaccination was not available but instead was widely ignored. Nonetheless, this result may offer hope for combating future variants of the rapidly changing virus as well as the dreaded breakthrough infections, in which severe outcomes have been seen in 10.5% of the vaccinated versus 26.5% of the unvaccinated group [177], with breakthrough even being fatal in 2% of cases [178].

Could a virus that is spreading so easily and is much deadlier than H1N1 influenza be kept under control if the human immune system could work at its fullest capacity? Zero mortality, a phrase used in the abstract, is of course an impossibility, as there is always a given intrinsic mortality rate for any age. Statistical variations in genetics as well as in lifestyle often prevent us from identifying the exact medical cause of death, especially when risk factors (i.e., comorbidities) and an acute infection are in competition with one another. Risk factors also tend to reinforce each other. In COVID-19, it is common knowledge that type II diabetes, obesity, and high blood pressure easily double the risk of death [160], depending on age. The discussion of whether a patient has died “because of” or “with” COVID-19 or “from” or only “with” his or her comorbidities thus seems obsolete. SARS-CoV-2 infection is statistically just adding to the overall mortality risk, but obviously to a much higher degree than most other infectious diseases or general risk factors.

The background section has shown that the vitamin D system plays a crucial role not only in the healthiness and strength of the skeletal system (rickets/osteoporosis) but also in the outcome of many infectious and/or autoimmune diseases [161,162]. Preexisting D3 deficiency is highly correlated in all of these previously mentioned cases.

Many argue that, because a *correlation does not imply causality*, a low D3 level may be merely a biomarker for an existing disease rather than its cause. However, the range of diseases for which existing empirical evidence shows an inverse relationship between disease severity and long-term D3 levels suggests that this assumption should be reversed [163].

This study investigated the correlation between vitamin D levels as a marker of a patient’s immune defense and resilience against COVID-19 and presumably other respiratory infections. It compared and merged data from two completely different datasets. The strength of the chosen approach lies in its diversity, as data from opposite and independent parts of the data universe yielded similar results. This result strengthens the hypothesis that a fatal outcome from COVID-19 infection, apart from other risk factors, is strongly dependent on the vitamin D status of the patient. The mathematical regressions suggested that the lower threshold for healthy vitamin D levels should lie at approximately 125 nmol/L or 50 ng/ml 25(OH)D3, which would save most lives, reducing the impact even for patients with various comorbidities.

This is – to our knowledge – the first study that aimed to determine an optimum D3 level to minimize COVID-19 mortality, as other studies typically limit themselves to identifying odds ratios for 2–3 patient cohorts split at 30 ng/ml or lower.

Another study confirmed that the number of infections clearly correlated with the respective D3 levels, with a cohort size close to 200,000 [115]. A minimum number of infections was observed at 55 ng/ml.

Does that mean that vitamin D protects people from getting infected? Physically, an infection occurs when viruses or bacteria intercept and enter body cells. Medically, infections are defined as having symptomatic aftereffects. However, a positive PCR test presumes the individual to be infectious even when there are no clinical symptoms and can be followed by quarantine. There is ample evidence that many people with a confirmed SARS-CoV-2 infection have not shown any symptoms [166].

A “physical infection”, which a PCR test can later detect, can only be avoided by physical measures such as disinfection, masks and/or virucidal sprays, which will prevent the virus from either entering the body or otherwise attaching to body cells to infect them. However, if we define “infection” as having to be clinically symptomatic, then we have to refer to it as “silent” to describe what happens when the immune system fights down the virus without showing any symptoms apart from producing specific T-cells or antibodies. Nevertheless, the PCR test will show such people as being “infected/infectious”, which justifies that they are counted as “cases” even without confirmation by clinical symptoms, e.g., in Worldometer Statistics [164].

Just as the D3 status correlates not only with the severity of symptoms but also with the length of the ongoing disease [165], it is fair to assume that the same reasoning also applies for silent infections. Thus, the duration in which a silent infection itself is active, i.e., infectious and will therefore produce a positive PCR result, may be reduced. We suggest that this may have a clear effect on the reproduction rate.

Thus, it seems clear that a good immune defense, be it naturally present because of good preconditioning or from an acquired cross immunity from earlier human coronavirus infections, cannot “protect” against the infection like physical measures but can protect against clinical symptoms. Finding only half as many “infected” patients (confirmed by PCR tests) with a vitamin D level >30 ng/ml [115] does not prove protection against physical infection but rather against its consequences – a reduction in the number of days of people being infectious must statistically lead to the demonstrated result of only half as many positive PCR tests recorded in the group >30 ng/ml vs. the group <30 ng/ml. This “protection” was most effective at ∼55 ng/ml, which agrees well with our results.

This result was also confirmed in a 2012 study, which showed that one of the fatal and most feared symptoms of COVID-19, the out-of-control inflammation leading to respiratory failure, is directly correlated with vitamin D levels. Cells incubated in 30 ng/ml vitamin D and above displayed a significantly reduced response to lipopolysaccharides (LPS), with the highest inflammatory inhibition observed at 50 ng/ml [167].

This result matches scientific data on the natural vitamin D3 levels seen among traditional hunter/gatherer lifestyles in a highly infectious environment, which were 110–125 nmol/L (45–50 ng/ml) [168].

There is a major discrepancy with the 30 ng/ml D3 value considered by the WHO as the threshold for sufficiency and the 20 ng/ml limit assumed by D-A-CH countries.

Three directors of Iranian Hospital Dubai also state from their practical experience that among 21 COVID-19 patients with D3 levels above 40 ng/ml (supplemented with D3 for up to nine years for ophthalmologic reasons), none remained hospitalized for over 4 days, with no cytokine storm, hypercoagulation or complement deregulation occurring [169].

Thus, we hypothesize that long-standing supplementation with D3 preceding acute infection will reduce the risk of a fatal outcome to practically nil and generally mitigate the course of the disease.

However, we have to point out that there are exceptions to that as a rule of nature: as in any multifactorial setting, we find a bell curve distribution in the activation of a huge number of genes that are under the control of vitamin D. There may be genetic reasons for this finding, but there are also additional influencing parameters necessary for the production of enzymes and cells of the immune system, such as magnesium, zinc, and selenium. Carlberg et al. found this bell curve distribution when verifying the activation of 500 - 700 genes contributing to the production of immune system-relevant cells and proteins after D3 supplementation [170]. Participants at the low end showed only 33% activation, while others at the high end showed well over 80% “of the 36 vitamin D3-triggered parameters”. Carlberg used the term (vitamin D3) low and high responders to describe what he saw.

This finding may explain why a “D3 deficient” high responder may show only mild or even no symptoms, while a low responder may experience a fatal outcome. It also explains why, on the one hand, many so-called “autoimmune” inflammation-based diseases do highly correlate with the D3 level based on, e.g., higher latitudes or higher age, when D3 production decreases, but why only parts of the population are affected: it is presumably the low responders who are mostly affected. Thus, for 68%-95% (1 or 2 sigma SDs), the suggested D3 level may be sufficient to fight everyday infections, and for the 2.5%-16% of high responders, it is more than sufficient and is completely harmless. However, for the 2.5%-16% of low responders, this level should be raised further to 75 ng/ml or even >100 ng/ml to achieve the same immune status as mid-level responders. A vitamin D3 test before the start of any supplementation in combination with the patient’s personal history of diseases might provide a good indication as to which group the patient belongs to and thus whether 50 ng/ml would be sufficient, or, if “normal” levels of D3 are found (between 20 and 30 ng/ml) along with any of the known D3-dependent autoimmune diseases, a higher level should be targeted as a precaution, especially as levels up to 120 ng/ml are declared to have no adverse effects by the WHO.

As future mutations of the SARS-CoV-2 virus may not be susceptible to the acquired immunity from vaccination or from a preceding infection, the entire population should raise their serum vitamin D level to a safe level as soon as possible. As long as enough vitamin K2 is provided, the suggested D3 levels are entirely safe to achieve by supplementation. However, the body is neither monothematic nor monocausal but a complicated system of dependencies and interactions of many different metabolites, hormones, vitamins, micronutrients, enzymes, etc. Selenium, magnesium, zinc and vitamins A and E should also be controlled for and supplemented where necessary to optimize the conditions for a well-functioning immune system.

A simple observational study could prove or disprove all of the above. If one were to test PCR-positive contacts of an infected person for D3 levels immediately, i.e., before the onset of any symptoms, and then follow them for 4 weeks and relate the course of their symptomatology to the D3 level, the same result as shown above must be obtained: a regression should cross the zero baseline at 45-55 ng/ml. Therefore, we strongly recommend the performance of such a study, which could be carried out with very little human and economic effort.

Even diseases caused by low vitamin D3 levels cannot be entirely resolved by ensuring a certain (fixed) D3 level for the population, as immune system activation varies. However, to fulfill Scribonius Largus’ still valid quote “primum non nocere, secundum cavere, tertium sanare” from 50 A.D., it should be the duty of the medical profession to closely look into a medication or supplementation that might help (tertium sanare) as long as it has no known risks (primum non nocere) within the limits of dosages that are needed for the blood level mentioned (secundum cavere).

Unfortunately, this does not imply that in the case of an acute SARS-CoV-2 infection, newly started supplementation with 25(OH)D3 will be a helpful remedy when calcidiol deficiency is evident, especially if this deficiency has been long lasting and caused or exacerbated typical comorbidities that can now aggravate the outcome of the infection. This was not a question we aimed to answer in this study.

### Limitations

This study does not question the vital role that vaccination will play in coping with the COVID-19 pandemic. Nor does it claim that in the case of an acute SARS-CoV-2 infection, a high boost of 25(OH)D3 is or could be a helpful remedy when vitamin D deficiency is evident, as this is another question. Furthermore, empirical data on COVID-19 mortality for vitamin D3 blood levels above 35 ng/ml are sparse.

## Conclusions

Although there are a vast number of publications supporting a correlation between the severity and death rate of SARS-CoV-2 infections and the blood level of vitamin D3, there is still an open debate about whether this relation is causal. This is because in most studies, the vitamin D level was determined several days after the onset of infection; therefore, a low vitamin D level may be the result and not the trigger of the course of infection.

In this publication, we used a meta-analysis of two independent sets of data. One analysis is based on the long-term average vitamin D3 levels documented for 19 countries. The second analysis is based on 1601 hospitalized patients, 784 who had their vitamin D levels measured within a day after admission, and 817 whose vitamin D levels were known pre-infection. Both datasets show a strong correlation between the death rate caused by SARS-CoV-2 and the vitamin D blood level. At a threshold level of 30 ng/ml, mortality decreases considerably. In addition, our analysis shows that the correlation for the combined datasets intersects the axis at approximately 50 ng/ml, which suggests that this vitamin D3 blood level may prevent any excess mortality. These findings are supported not only by a large infection study, showing the same optimum, but also by the natural levels observed in traditional people living in the region where humanity originated from that were able to fight down most (not all) infections in most (not all) individuals.

Vaccination is and will be an important keystone in our fight against SARS-CoV-2. However, current data clearly show that vaccination alone cannot prevent all SARS-CoV-2 infections and dissemination of the virus. This scenario possibly will become much worse in the case of new virus mutations that are not very susceptible to the current vaccines or even not sensitive to any vaccine.

Therefore, based on our data, the authors strongly recommend combining vaccination with routine strengthening of the immune system of the whole population by vitamin D3 supplementation to consistently guarantee blood levels above 50 ng/ml (125 nmol/l). From a medical point of view, this will not only save many lives but also increase the success of vaccination. From a social and political point of view, it will lower the need for further contact restrictions and lockdowns. From an economical point of view, it will save billions of dollars worldwide, as vitamin D3 is inexpensive and – together with vaccines – provides a good opportunity to get the spread of SARS-CoV-2 under control.

Although there exists very broad data-based support for the protective effect of vitamin D against severe SARS-CoV-2 infections, we strongly recommend initiating well-designed observational studies as mentioned and/or double-blind randomized controlled trials (RCTs) to convince the medical community and the health authorities that vitamin D testing and supplementation are needed to avoid fatal breakthrough infections and to be prepared for new dangerous mutations.

## Data Availability

The datasets generated and/or analyzed during the current study have been made available online.

https://doi.org/10.7910/DVN/7FSWNL

## Declarations

### Ethics approval and consent to participate

Not applicable.

### Consent for publication

Not applicable.

### Availability of data and materials

The datasets generated and/or analyzed during the current study have been made available online [171].

## Authors’ Information

### Affiliations

None.

### Contributions

Conceptualization: L.B.

Data curation: L.B. and J.V.M.

Writing – Background: B.G.

Writing – Methods and Results: J.V.M.

Writing – Discussion: L.B.

Writing – Abstract/Conclusion/review & editing: L.B., B.G., and J.V.M.

## Acknowledgments

This manuscript was edited for English language by American Journal Experts (AJE).

